# Evaluation of smartphone-based cough data in amyotrophic lateral sclerosis as a potential predictor of functional disability

**DOI:** 10.1101/2024.03.24.24304803

**Authors:** Pedro Santos-Rocha, Nuno Bento, Duarte Folgado, André Valério Carreiro, Miguel Oliveira Santos, Mamede de Carvalho, Bruno Miranda

## Abstract

**Objectives:** Cough dysfunction is a feature of patients with amyotrophic lateral sclerosis (ALS). The cough sounds carry information about the respiratory system and bulbar involvement. Our goal was to explore the association between cough sound characteristics and the respiratory and bulbar functions in ALS.

**Methods:** This was a single-center, cross-sectional, and case-control study. On-demand coughs from ALS patients and healthy controls were collected with a smartphone. A total of 31 sound features were extracted for each cough recording using time-frequency signal processing analysis. Logistic regression was applied to test the differences between patients and controls, and in patients with bulbar and respiratory impairment. Support vector machines (SVM) were employed to estimate the accuracy of classifying between patients and controls and between patients with bulbar and respiratory impairment. Multiple linear regressions were applied to examine correlations between cough sound features and clinical variables.

**Results:** Sixty ALS patients (28 with bulbar dysfunction, and 25 with respiratory dysfunction) and forty age- and gender-matched controls were recruited. Our results revealed clear differences between patients and controls, particularly within the frequency-related group of features (AUC 0.85, CI 0.79- 0.91). Similar results were observed when comparing patients with and without bulbar dysfunction; and with and without respiratory dysfunction. Sound features related to intensity displayed the strongest correlation with disease severity.

**Discussion:** We found a good relationship between specific cough sound features and clinical variables related to ALS functional disability. The findings relate well with some expected impact from ALS on both respiratory and bulbar contributions to the physiology of cough. Finally, our approach could be relevant for clinical practice, and it also facilitates home-based data collection.

## 1. Introduction

Amyotrophic Lateral Sclerosis (ALS) is a progressive neurodegenerative disease characterized by the loss of both upper and lower motor neurons (1). The consequent motor dysfunction leads to symptoms affecting limbs, bulbar and respiratory muscles – and eventual death by respiratory insufficiency or infection (2). In general, the disease is characterized by significant variability in onset region, as well as in the pattern and rate of progression (3–7).

The majority of cases show either a spinal phenotype or a bulbar variant, but some patients present initial trunk or respiratory involvement (5,8,9). However, an early respiratory and bulbar impairment are associated with poor quality of life, malnutrition, and early mortality (10,11). Non-invasive pulmonary function tests, in particular forced vital capacity (FVC), has long been used for respiratory assessment and monitoring. However, they require cooperative patients, good lips strength, and repeated testing to ensure consistency of measurements (12). On the other hand, an established and comprehensive clinical scale to objectively monitor bulbar disease and respiratory progression in ALS has yet to be achieved (13).

During the last few years, objective evaluation of cough sounds, in particular evaluating its quantitative characteristics in terms of sound frequency or intensity, has gained popularity for detecting and distinguishing different respiratory dysfunctions (12,14–18). The increasing evidence concerning the objective evaluation of cough is also grounded by the physiological mechanisms of coughing which require considerable coordination and timing of breathing, thus being sensitive to abnormalities in the respiratory system (19,20). Physiologically, cough involves a deep inspiration, followed by vigorous contraction of the expiratory muscles (in particular the abdominal muscles) against a closed glottis. When a certain subglottic pressure is reached, the glottis opens, producing one initial supramaximal expiratory airflow followed by a longer-lasting lower expiratory flow, generating the cough sound at the same time. Importantly, such physiological mechanisms for a normal cough also rely on a normal bulbar function, being especially relevant for the glottis and intrinsic laryngeal muscles’ performance. The latter muscles are the ones responsible for the dimensions of the glottis rhyme (i.e., the tension regulation of the vocal ligaments) and changes in laryngeal opening and closing – which are key properties of the cough sound. In more advanced ALS, cough is generally weak and absent (21,22); this causes inability to clear secretions, eases choking and impairs protection of the respiratory system – often leading to aspiration pneumonia. Recent progress has been made to take advantage of sensors to monitor the functional state of ALS patients, including for home-based assessments (23–26). Stegmann et al. (27) used a mobile application (app) installed on the patient’s mobile device to record speech acoustics and to predict their forced vital capacity (FVC). Furthermore, Vashkevich et al. (28) proposed an approach to voice assessment for automatic systems to differentiate healthy individuals from ALS patients (based on sustained phonation of the vowels /a/ and /i/). They used a wide range of acoustic features to achieve high accuracy in this classification. A feasibility study utilizing cough sound to differentiate between healthy individuals and those with ALS was recently conducted by Cebola et al. (29). The study endorsed the viability of using coughs for remote monitoring; however, the sample size was limited and not gender-matched. Despite previous efforts focused on studying speech and cough acoustics, very few studies have comprehensively explored the potential of cough sound analysis in ALS.

In this study, we hypothesize that cough sound features obtained by a smartphone and using time- and frequency-domain analysis, could inform about bulbar and respiratory impairments in ALS patients. Thus, the present work aims to: 1) evaluate if the sound features of a voluntary cough in ALS patients are different from age- and gender-matched healthy controls; 2) correlate cough sound features with functional status, respiratory and bulbar impairment in ALS patients; and 3) test the hypothesis that frequency sound features have a stronger association with bulbar dysfunction, while intensity sound features are more closely related to respiratory dysfunction. Furthermore, we aimed to evaluate the usefulness of machine learning for conducting future home-based assessments, by recording audio samples with a commonly available device, in an ecological setting.

## 2. Materials and Methods

### 2.1 Study design and participants

This was a single-center, cross-sectional, case-control study that was part of a broader ALS project (HomeSenseALS - PTDC/MEC-NEU/6855/2020). We included consecutive ALS patients according to Gold Coast criteria (30). All patients were followed at our ALS clinic in Lisbon, and had full neurological, neurophysiological, neuroimaging and blood tests to rule out mimicking conditions (31). Patients with a previous history of lung disorders, with resting dyspnea, severe cognitive involvement impairing the understanding of the voluntary coughing task, and those declining to participate were excluded. In the control group we included healthy age- and gender-matched controls (in general spouses of the ALS patients and people working in the institution). The recruitment started on April 4, 2022, and was concluded on August 31, 2023. The study was approved by the local research ethics committee of the Centro Académico de Medicina de Lisboa (CAML-Ref. 146/21). All participants gave written informed consent, which was in accordance with the declaration of Helsinki.

### 2.2 Clinical evaluation

For ALS patients, we collected demographic data including age, sex, body mass index (BMI), smoking habits, disease duration, and the region of disease onset. To evaluate the functional disability, we used the revised functional ALS rating scale (ALSFRS-R) (32).

Respiratory symptoms were determined based on the ALSFRS-R respiratory subscore (which consists of questions 10 through 12 pertaining to dyspnea, orthopnea, and respiratory insufficiency); patients with a score less than 12 were considered to have respiratory dysfunction. Sitting predicted FVC (FVC%) was measured using a computer-based USB spirometer (microQuark®, Cosmed®), the best of three reliable maneuvers was used for statistics (11). In addition to FVC%, the following respiratory measures were also included: maximum expiratory and inspiratory pressures (MIP% and MEP%, respectively) and cough peak flow (CPF). Similarly, bulbar symptoms were evaluated using the ALSFRS-R bulbar subscore (which consists of questions 1 through 3 about speech, salivation, and swallowing). Patients with a score less than 12 were considered having bulbar dysfunction. This data was accessed retrospectively, between August 31, 2023, and October 31, 2023. The authors had no access to information that could identify individual participants during or after data collection.

### 2.3 Cough sound: recording, signal processing and feature extraction

All subjects were instructed to perform, while seated in a quiet room, three voluntary coughs (to ensure a repeatable sound relationship). The sound recordings were done using a smartphone, placed approximately 20-25 cm away from the mouth and at an angle of approximately 45° (as described in (12)). These procedures aimed to remove effects of wind noise produced when one rapid expulsion of air directly hits the microphone. For patients, the cough sounds were recorded during a routine patient’s clinical visit, after ensuring that the patient was resting for a period longer than 10 minutes, and comfortable without dyspnea.

After the cough data collection, the raw signal was processed with Librosa – a Python package for audio signal analysis (33). The analysis was conducted using a frame length of 2048 samples per frame and a hop length of 512. In order to minimize potential biases stemming from the beginning and end of the recordings (and to ensure that the analysis was focused solely on the cough time frames) the split function of Librosa was employed with a cutoff of 20 decibels eliminating the initial and final periods of silence in the cough recordings.

Once the pre-processing was completed, the generated cough sound signals were analyzed to extract audio-based features. For this, we used the *Time Series Feature Extraction Library* (TSFEL) that automatically extracts over 60 different features on the statistical, temporal, and spectral domains (34).

In light of prior research findings (12,29,35,36) and relevance in general sound analysis, we pre-selected 11 features based on the time domain and 20 features based on the frequency domain (see details in **Table 1**). To enhance the interpretation of the results, we subsequently categorized these features into three distinct groups, each pertaining to specific underlying information (with potential relevance for the various physiological steps of coughing): 1) a group encompassing sound frequency-related features – the frequency group; 2) another group comprising sound intensity-related features – the intensity group; and 3) a final group that combines features of both frequency and intensity domains – the mixed group. All extracted features were normalized to their maximum value (with a range between -1 and 1) (**Fig 1**).

**Figure 1.**
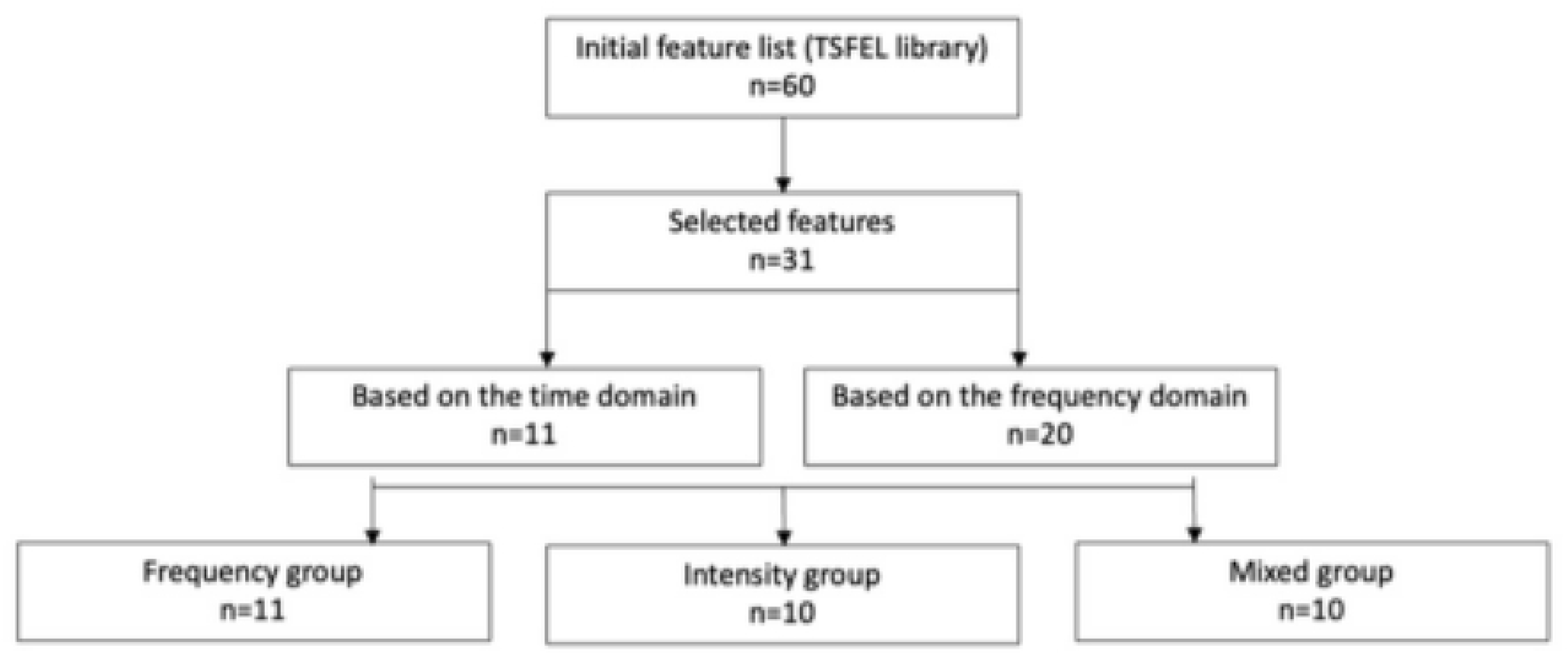
The workflow of feature categorization according to frequency, intensity, and mixed groups.

**Table 1.**
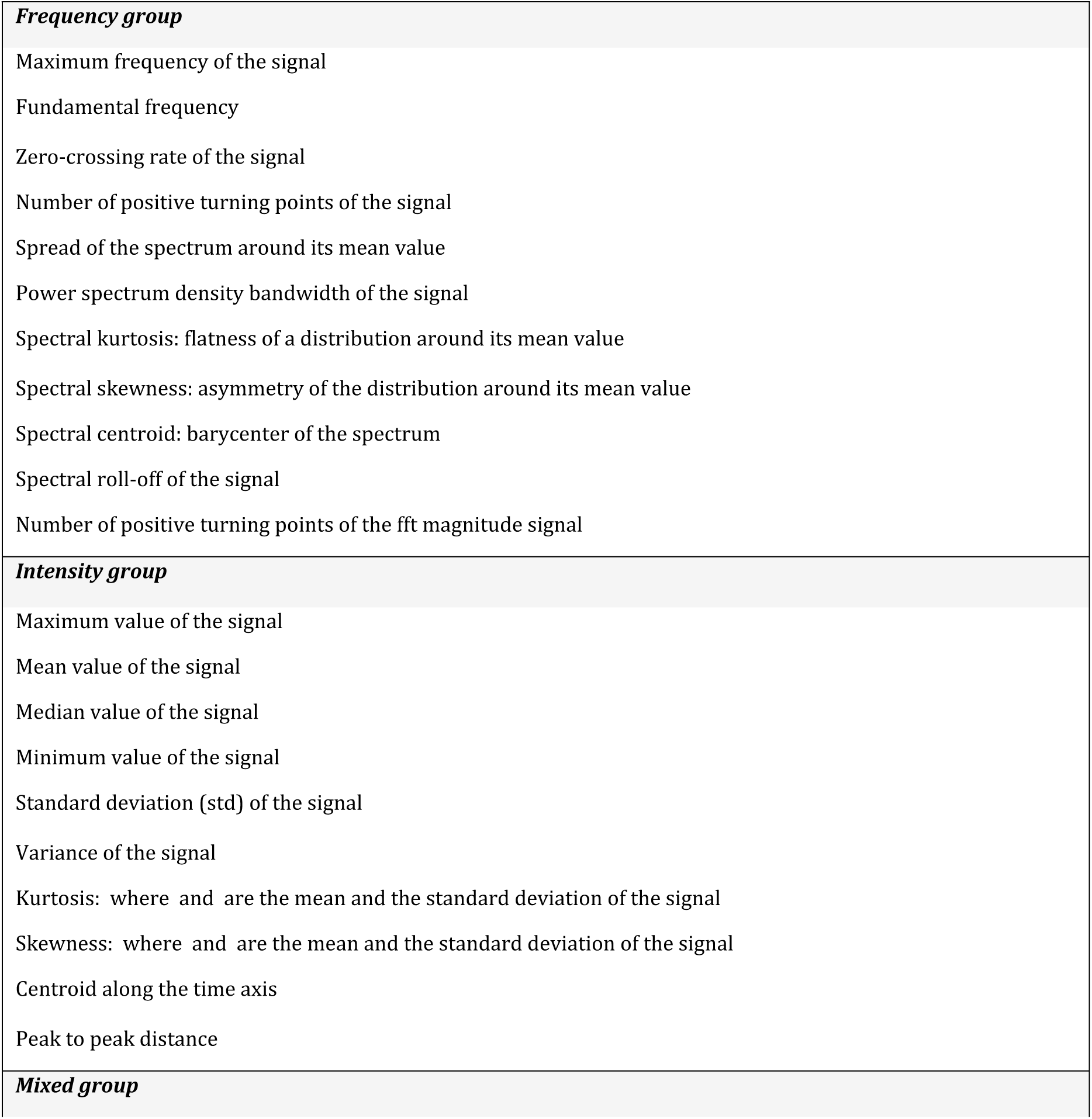
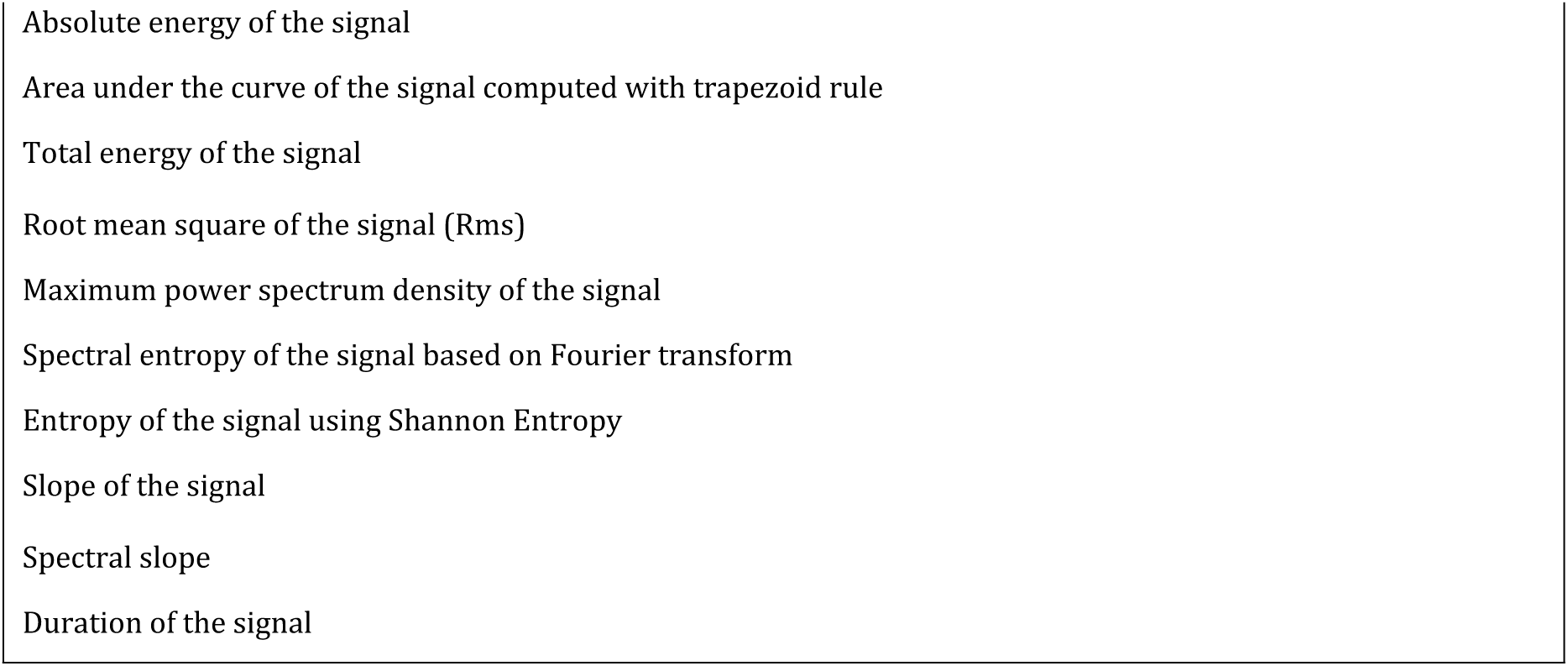
List of cough sound features subjected to analysis.

| ***Frequency group*** |
| --- |
| Maximum frequency of the signal |
| Fundamental frequency |
| Zero-crossing rate of the signal |
| Number of positive turning points of the signal |
| Spread of the spectrum around its mean value |
| Power spectrum density bandwidth of the signal |
| Spectral kurtosis: flatness of a distribution around its mean value |
| Spectral skewness: asymmetry of the distribution around its mean value |
| Spectral centroid: barycenter of the spectrum |
| Spectral roll-off of the signal |
| Number of positive turning points of the fft magnitude signal |
| ***Intensity group*** |
| Maximum value of the signal |
| Mean value of the signal |
| Median value of the signal |
| Minimum value of the signal |
| Standard deviation (std) of the signal |
| Variance of the signal |
| Kurtosis: where $\mu$ and $\sigma$ are the mean and the standard deviation of the signal |
| Skewness: where $\mu$ and $\sigma$ are the mean and the standard deviation of the signal |
| Centroid along the time axis |
| Peak to peak distance |
| ***Mixed group*** |
| Absolute energy of the signal |
| Area under the curve of the signal computed with trapezoid rule |
| Total energy of the signal |
| Root mean square of the signal (Rms) |
| Maximum power spectrum density of the signal |
| Spectral entropy of the signal based on Fourier transform |
| Entropy of the signal using Shannon Entropy |
| Slope of the signal |
| Spectral slope |
| Duration of the signal |

### 2.4 Machine-learning analysis

After the feature extraction step, a dataset was built for the purpose of a binary classification task with the objective of distinguishing between ALS patients and control subjects, patients with and without bulbar dysfunction, and patients with and without respiratory dysfunction. The dataset was partitioned, with 75% of the data allocated to the training set and the remaining data designated for testing. The process of shuffling resulted in a well-balanced test set in terms of class, age, gender, and dataset distribution. To identify a small subset of relevant features for the objective analysis of bulbar and respiratory ALS dysfunction, the extracted cough sound features underwent feature selection using the sequential feature selection (SFS) algorithm based on a logistic regression (LR) classifier. Through SFS, we selected the cough sound features that were strongly correlated with the class, thus removing the less relevant features from the original dataset. Subsequently, the main classification task was performed by training a support vector machine (SVM) classifier based on the linear kernel. In this process, only the most relevant features, which were selected in the preceding step were considered, in an attempt to reduce the potential for overfitting. For model evaluation, ROC-AUC (area under the curve) scores were calculated over five iterations, each with a distinct random seed, so that it would be possible to estimate the 95% confidence interval. This comprehensive procedure facilitated the assessment of model stability and reliability.

### 2.5 Statistical analysis

Data analysis was performed using Python version 3.11.2 (Python Software Foundation). For the significance level, *α*=0.05 was considered. Descriptive statistics consisted of frequencies (with proportions) for categorical variables and mean values (with standard deviation) for continuous variables. To compare mean values, parametric tests such as the two-sample t-test or the one-way ANOVA were applied. If the normality assumption of the continuous variable was violated (significant Kolmogorov-Smirnov test with an absolute skewness > 2), non-parametric tests such as Mann-Whitney U-test or Kruskal-Wallis test were considered and results reported, if different from parametric analysis.

ROC analyses were performed to identify the ROC-AUC of the SVM, for discriminating between:

(1) controls vs. ALS – with the frequency group of features;
(2) controls vs. ALS – with the intensity group of features;
(3) controls vs. ALS – with the mixed group of features.

Similar analyses were carried out for the comparison between patients with bulbar dysfunction vs. those without; and for patients with respiratory dysfunction vs. those without. Age and gender were added into each set of features, enabling the SVM model to consider these important demographic factors.

Finally, we examined how each of the selected features related to the disability score and pulmonary function tests in ALS patients. For the former, multiple linear regression models were used, having the ALSFRS-R total score as dependent variable and age and gender as confounding variables. On the other hand, simple linear correlations were used to elucidate associations between sound features and pulmonary function measurements, including FVC%, MEP%, MIP% and CPF.

## 3. Results

### 3.1 Demographics and clinical characteristics

We analyzed 300 cough sounds recordings from a total of 100 subjects (60 ALS patients and 40 controls - 3 cough sounds each). The demographic and clinical characteristics of participants are shown in **Table 2**. Groups had no significant differences in terms of age (p= 0.79) or sex distribution (p= 0.21). There were no statistically significant differences between patients with vs without respiratory dysfunction, as well as between patients with vs without bulbar dysfunction, in terms of age, BMI, disease duration and percentage of smokers (all t-tests with p- values > 0.05). However, the frequency of females was higher in the group with bulbar dysfunction (71% vs 34%, p< 0.001) and in the group of patients with respiratory dysfunction (72% vs 43%, p< 0.05).

**Table 2.** Baseline characteristics of whole ALS patients population (n=60), and controls (N=40).

| CLINICAL CHARACTERISTICS | ALS PATIENTS<br>N=60 | CONTROLS<br>N=40 |
| --- | --- | --- |
| AGE (mean±SD) | 61.87 ± 11.4 | 62.58 ± 15.63 |
| **GENDER** |  |  |
| MEN | 27 (45%) | 13 (32.5%) |
| WOMAN | 33 (55%) | 27 (64.5%) |
| **BMI (KG/M<sup>2</sup>) (mean±SD)** | 26.3 ± 4.1 | |
| **SYMPTOM DURATION (MONTHS)** |  |  |
| MEDIAN | 25 |  |
| 1<sup>ST</sup>-3<sup>RD</sup> INTERQUATILE RANGE | 6 – 141 |  |
| **DISEASE ONSET** |  |  |
| BULBAR ONSET | 15 (25%) |  |
| UPPER LIMB ONSET | 17 (28.3%) |  |
| LOWER LIMB ONSET | 28 (46.7%) |  |
| **ALSFRS-R TOTAL SCORE (0-48) (mean±SD)** | 34.9 (7.8) |  |
| **BULBAR DYSFUNCTION** | 28 (46.7%) |  |
| **RESPIRATORY DYSFUNCTION** | 25 (41.7%) |  |

BMI, body mass index; ALSFRS-R, revised amyotrophic lateral sclerosis functional rating scale.

### 3.2 Cough sound features in ALS and healthy controls

We started by comparing the frequency group of cough sound features in ALS patients vs. controls. The SFS algorithm selected six features of the thirteen initially proposed, including fundamental frequency, number of the spectrum positive turning points, spectral bandwidth, spectral roll-off, spectral dispersion, and zero-crossing rate (ZCR). Following ROC analysis (**Fig 2**), the prediction ROC-AUC of the final model with the seven selected features was 0.85 (IC 95%: 0.79-0.91).

**Figure 2.**
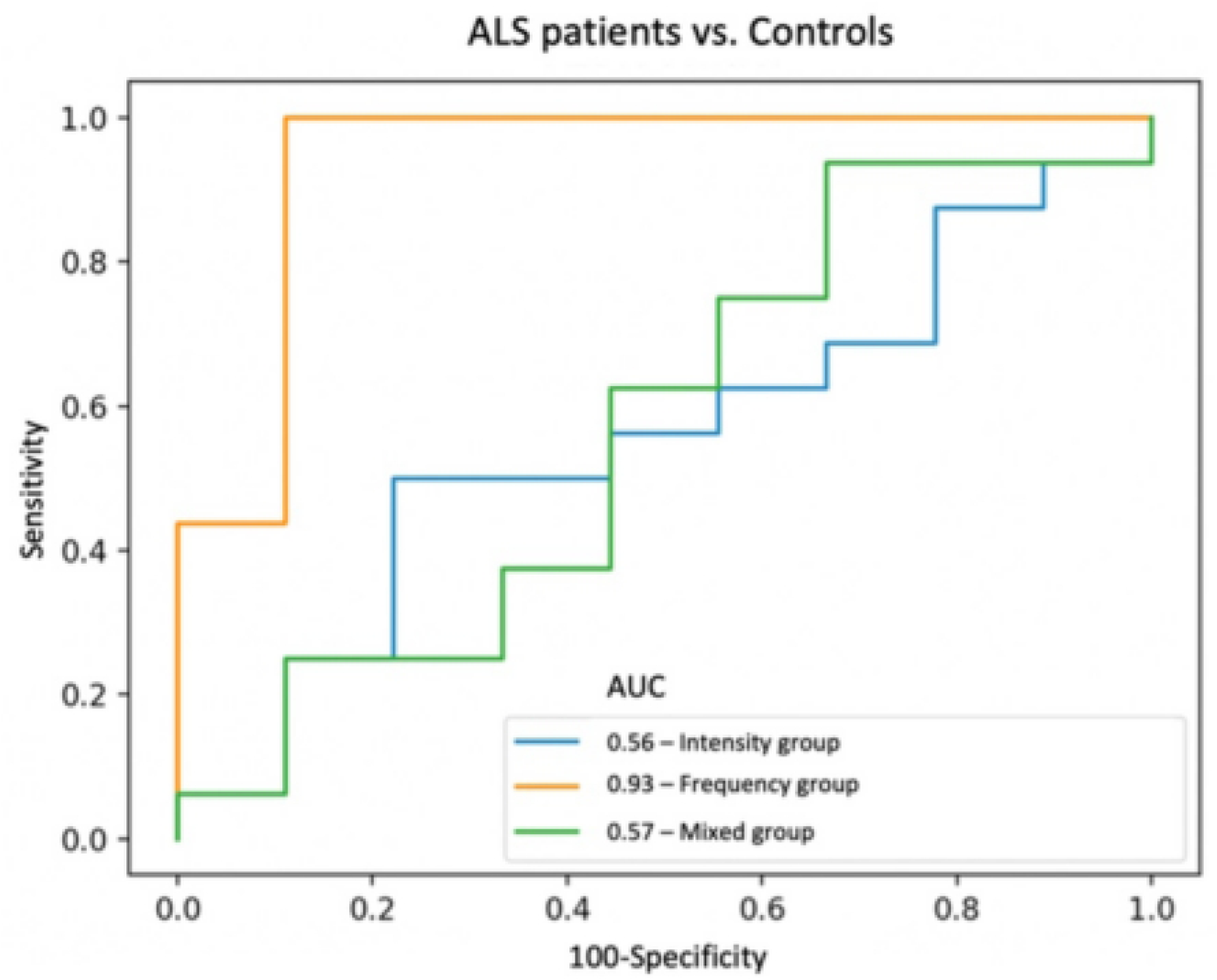
Support vector machine (SVM) analysis of all cough sound samples. Receiver operating characteristic curves (ROC) were calculated with a SVM to differentiate ALS patients and controls for the three different groups of cough sound features. One of the five model’s iterations is demonstrated (settings: k_features = ‘best’; forward = ‘False’; scoring = ‘accuracy’; cv = ‘5’; random_state = ‘41’).

Similar analyses were performed for the remaining intensity and mixed groups. The SFS applied to the intensity group of features resulted in the selection of four features out of the initial twelve. Specifically, these included the temporal centroid, the mean, and the kurtosis of the signal, and presumed gender. However, the model with the four intensity features exhibited a modest performance of 0.59 (IC 95%: 0.52-0.66). Also, to note that only the temporal centroid and the kurtosis of the signal demonstrated significant discriminative capability between an ALS-related cough and a control cough.

Regarding the mixed group, the model comprised the following selected features: absolute energy, spectral and time entropies, maximum power of the signal, total amount of energy, and presumed gender. However, similar to the intensity group, the overall model performance was only modest, yielding an ROC-AUC of 0.6 (IC 95%: 0.52-0.68). To also note that only the time and spectral entropies exhibited significant predictive capability for distinguishing between ALS patients and healthy controls. **Fig 3** shows an example where it is evident that the primary distinctions between an ALS and a control cough lie within the frequency group of features. **Table 3** shows all statistical values.

**Figure 3.**
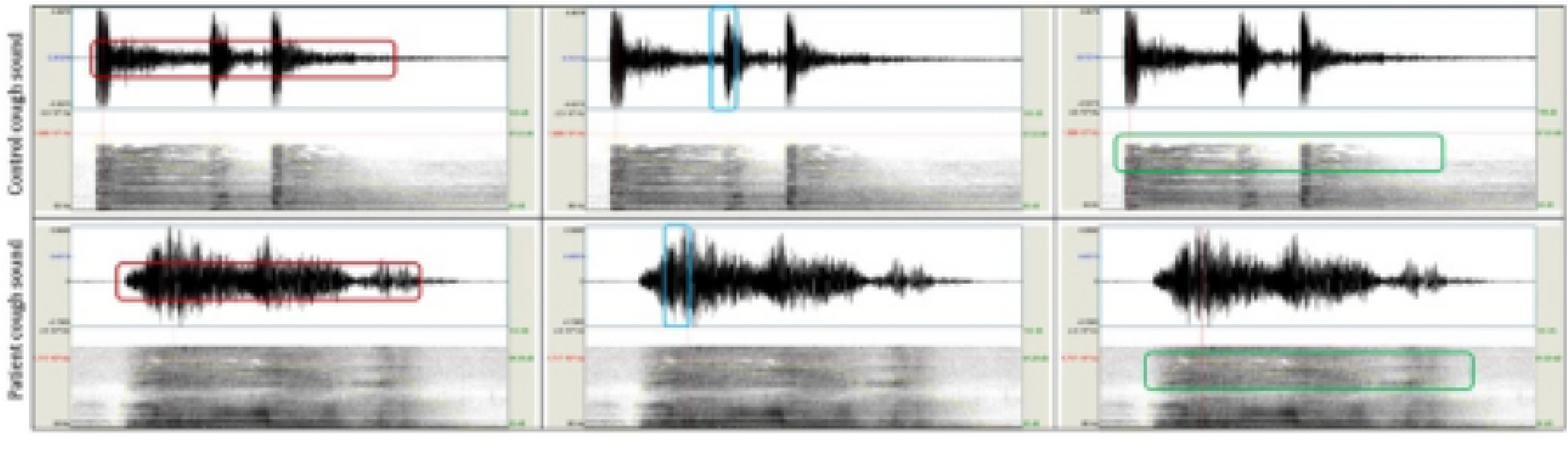
An example of the analysis of sound waves in voluntary coughing of a healthy control (upper row) and an ALS patient (lower row). Each column of the image depicts different features of the three distinct groups of features. The first column highlights the frequency features associated with the repetition rate of one event, such as the number of times that the signal passes the zero line or the number of positive turning points. The second column emphasizes the intensity features, such as the signal amplitude or peak distance. Lastly, the third column shows features that provide information on both frequency and intensity, such as the signal power and entropy. The main differences were observed in the frequency group. To note that these cough signals did not undergo pre-processing procedures.

**Table 3.** F values from regression analyses contributing of Control vs. ALS classification to performance on each voice sound variable.

| GROUP | FEATURES | F value | P value |
| --- | --- | --- | --- |
| Frequency group | Fundamental frequency | 9.19** | 0.0031 |
|  | Number of the spectrum positive turning points | 2.02 | 0.16 |
|  | Spectral bandwidth | 8.60** | 0.0042 |
|  | Spectral roll-off | 4.38* | 0.039 |
|  | Spectral dispersion | 10.23** | 0.0019 |
|  | ZCR | 20.19*** | < 0.001 |
| Intensity group | Temporal centroid | 11.55*** | < 0.001 |
|  | Mean amplitude of the signal | 0.54 | 0.47 |
|  | Kurtosis | 9.90** | 0.0022 |
| Mixed group | Absolute energy | 0.42 | 0.52 |
|  | Entropy | 7.25** | 0.0083 |
|  | Spectral entropy | 10.71** | 0.0015 |
|  | Maximum power of the signal | 0.39 | 0.53 |
|  | Total amount of energy | 1.03 | 0.31 |

Correlation is significant at the 0.05 level \*; Correlation is significant at the 0.01 level \*\*; Correlation is significant at the 0.001 level \*\*\*;

### 3.3 Correlations with the overall functional disability

Next, we started to focus more specifically on the ALS patients, and how the cough sound features were related to the functional disability of the disease. We found that the intensity, as well as the mixed group features, exhibited the strongest correlations with ALSFRS-R total score – indicating that patients with more severe symptoms produced cough sounds with a greater relative impact on the intensity domain (**Fig 4**). Broadly, patients in more advanced functional states produce less intense cough sounds.

**Figure 4.**
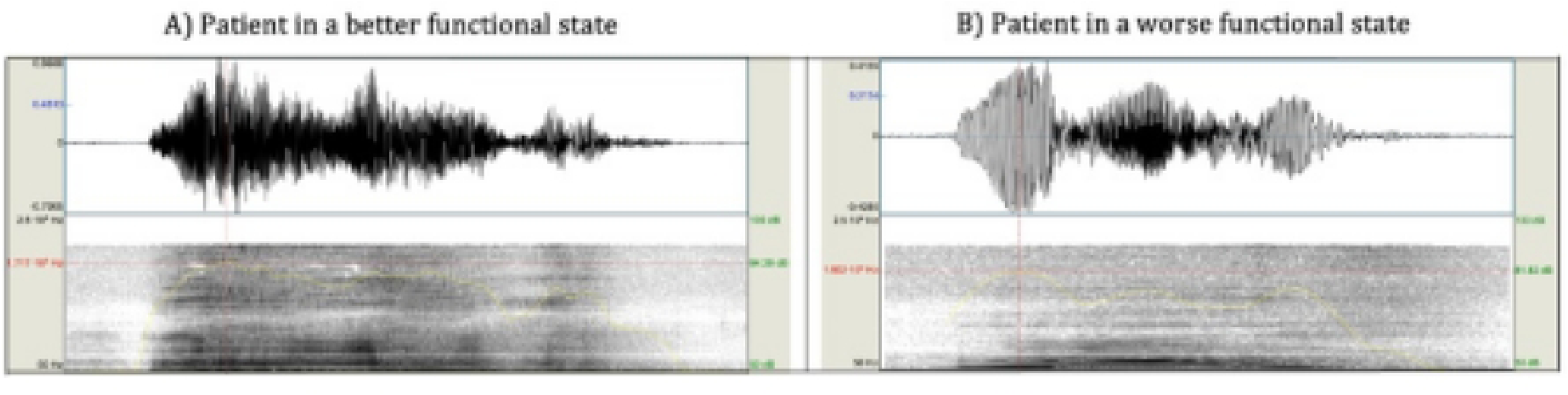
Analysis of sound waves in voluntary coughing: comparison between patients in different disease states. The left image represents the cough sound of one patient in a better functional state (Female; 60> years old; ALSFRS-R total score of 39) versus the right image, which represents the cough sound of one patient in a worse functional state (Female; 60> years old; ALSFRS-R total score of 20). The main differences are presented in the intensity-related group of features. (Signal without pre-processing)

Moderate but significant correlations have been found between the ALSFRS-R total score and various intensity features, including the maximum amplitude (beta= 0.43, p= 6.85e-4), the standard deviation of the amplitude of the signal (beta= 0.33, p= 1.55e-5) and the peak-to-peak distance (beta= 0.44, p= 4.07e-4). Moreover, we found moderate to strong negatively significant correlations, with the maximum cough sound power (beta= -0.58, p= 2.54e-10); and moderate positively significant correlations with the area under the curve of the signal and the absolute energy (beta= 0.35, p= 9.76e-6; beta= 0.34, p= 1.67e-6, respectively).

Despite being effective in distinguishing cough sounds from ALS and healthy controls, the frequency group of features showed weaker associations with the functional status of the disease. Our analysis revealed that ZCR (beta= 0.20, p= 3.01e-6), and the number of positive turning points (beta= 0.25, p= 1.10e-5) exhibited weak positively significant correlations with the ALSFRS-R total score. In contrast, the spectral centroid (beta= 0.38, p= 1.97e-4), and spectral bandwidth (beta= 0.32, p= 8.83e-8) demonstrated stronger correlations with the functional state of the disease, making them the best-correlated features in this frequency group (see **Table 4**).

**Table 4.** Correlations between the ALSFRS-R total score and different cough sound features. Results are adjusted for age and gender.

| GROUP | FEATURES | METRICS |  |  |
| --- | --- | --- | --- | --- |
| | | $\beta$ | T VALUE | P VALUE |
| **Intensity**<br>0.59 (IC 95%: 0.52-0.66)<br>model ROC-AUC | Maximum amplitude | 0.43 | 3.59 | <0.001 |
|  | Standard deviation of the signal | 0.33 | 4.73 | <0.001 |
|  | Peak distance | 0.44 | 3.76 | <0.001 |
| **Mixed**<br>0.60 (IC 95%: 0.52-0.68)<br>model ROC-AUC | Maximum power | -0.58 | 7.68 | <0.001 |
|  | Area under the curve | 0.35 | 4.86 | <0.001 |
|  | Absolute energy | 0.34 | 5.35 | <0.001 |
| **Frequency**<br>0.85 (IC 95%: 0.79-0.91)<br>model ROC-AUC | Zero-crossing rate | 0.20 | 5.19 | <0.001 |
|  | Spectral centroid | 0.38 | 3.98 | <0.001 |
|  | Number of positive turning points | 0.25 | 4.83 | <0.001 |
|  | Spectrum bandwidth | 0.32 | 6.14 | <0.001 |

### 3.4 Differences between ALS patients with and without respiratory dysfunction

We identified both intensity and mixed groups of features as the most effective in distinguishing patients with respiratory dysfunction from those without (with positive associations). In terms of intensity-related features, the predictors that remained in the final model were the maximum and standard deviation of the signal as well as peak-to-peak distance, yielding a prediction ROC-AUC of 0.67 (IC 95%: 0.56-0.78).

Regarding the mixed group features, the SFS application resulted in a final model comprising maximum power, spectral entropy, and the spectral slope. The prediction ROC-AUC of this model was 0.65 (IC 95%: 0.55-0.75).

Finally, the group of frequency-related features included in the final model the spectral bandwidth and centroid, the spectral skewness, and the ZCR; and yielding a prediction ROC-AUC of 0.59 (IC 95%: 0.49-0.69). **Table 5** shows all statistical values.

**Table 5.**
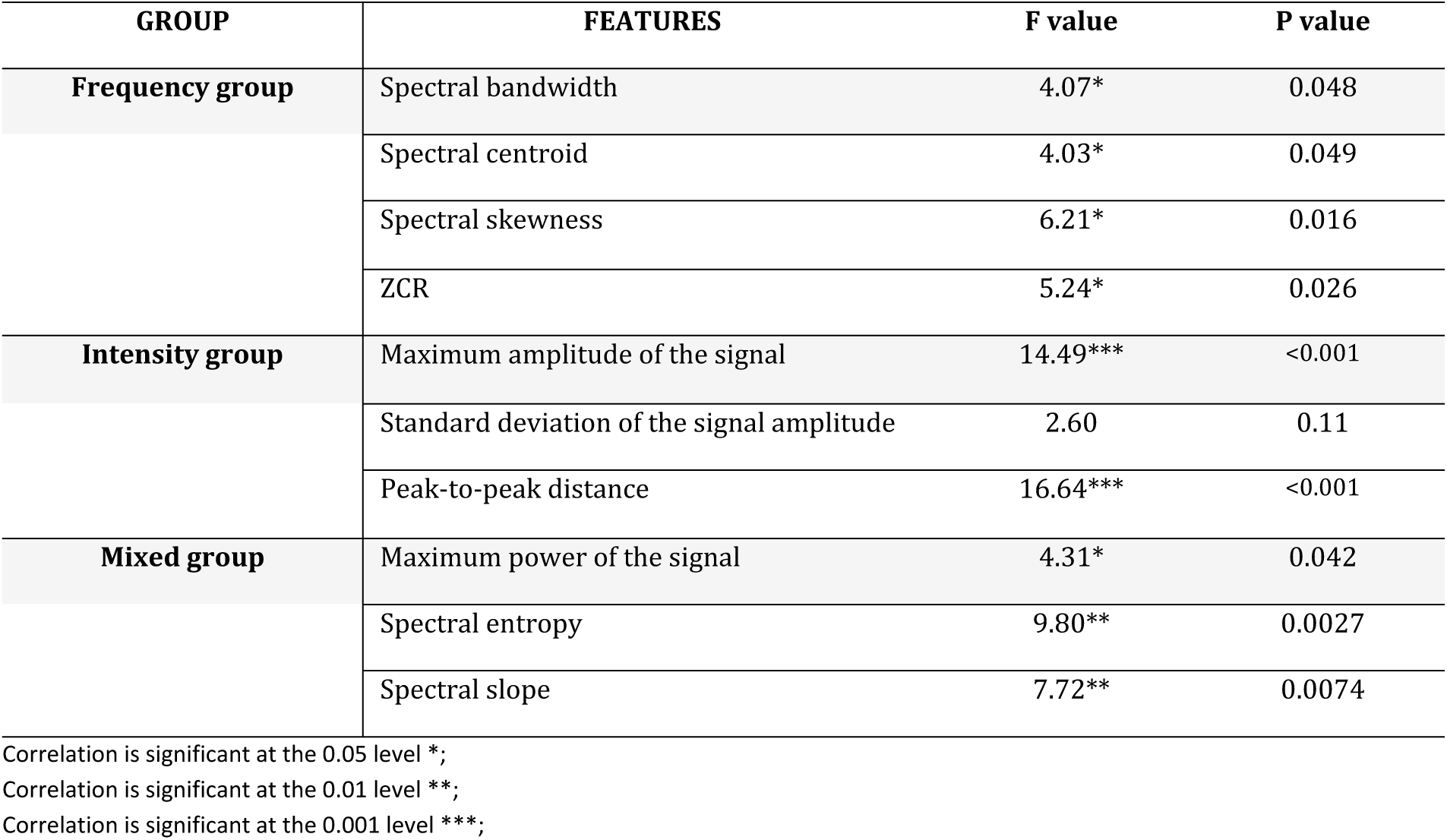
F values from regression analyses contributing of with respiratory vs. without respiratory dysfunction classification to performance on each voice sound variable.

| GROUP | FEATURES | F value | P value |
| --- | --- | --- | --- |
| Frequency group | Spectral bandwidth | 4.07* | 0.048 |
|  | Spectral centroid | 4.03* | 0.049 |
|  | Spectral skewness | 6.21* | 0.016 |
|  | ZCR | 5.24* | 0.026 |
| Intensity group | Maximum amplitude of the signal | 14.49*** | <0.001 |
|  | Standard deviation of the signal amplitude | 2.60 | 0.11 |
|  | Peak-to-peak distance | 16.64*** | <0.001 |
| Mixed group | Maximum power of the signal | 4.31* | 0.042 |
|  | Spectral entropy | 9.80** | 0.0027 |
|  | Spectral slope | 7.72** | 0.0074 |

Correlation is significant at the 0.05 level \*; Correlation is significant at the 0.01 level \*\*; Correlation is significant at the 0.001 level \*\*\*;

#### 3.4.1 Correlations between cough sound analysis and respiratory function assessments

Out of the complete cohort of 60 ALS patients, 47 had respiratory function testing at the time of cough sound data recording (< 6 weeks). In this group, mean ALSFRS-R was 38, and the average FVC%, MIP%, MEP%, and CPF were 77% (18.33 SD), 93.6% (32.8 SD), 79.8% (27 SD), and 288.8 L/min (109 SD), respectively.

**Table 6** shows that the intensity and mixed groups of features exhibited a moderate (negative) significant correlations with FVC%. However, no significant correlations were observed for CPF, MIP%, and MEP%.

**Table 6.**
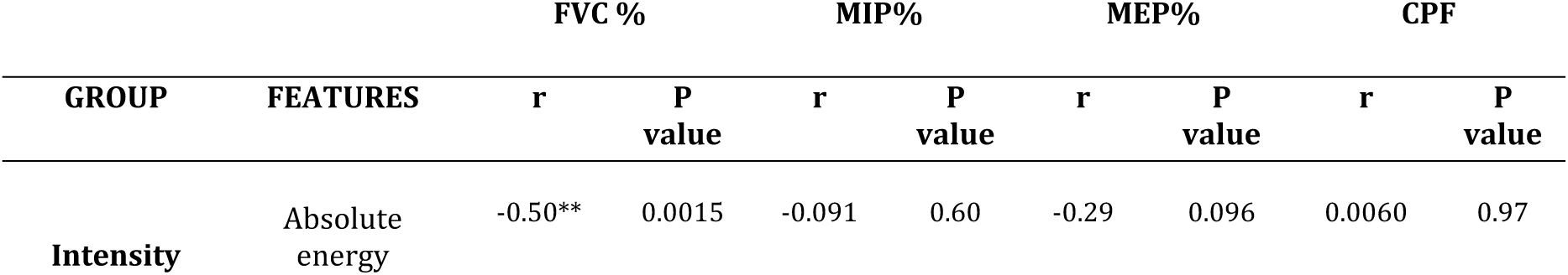
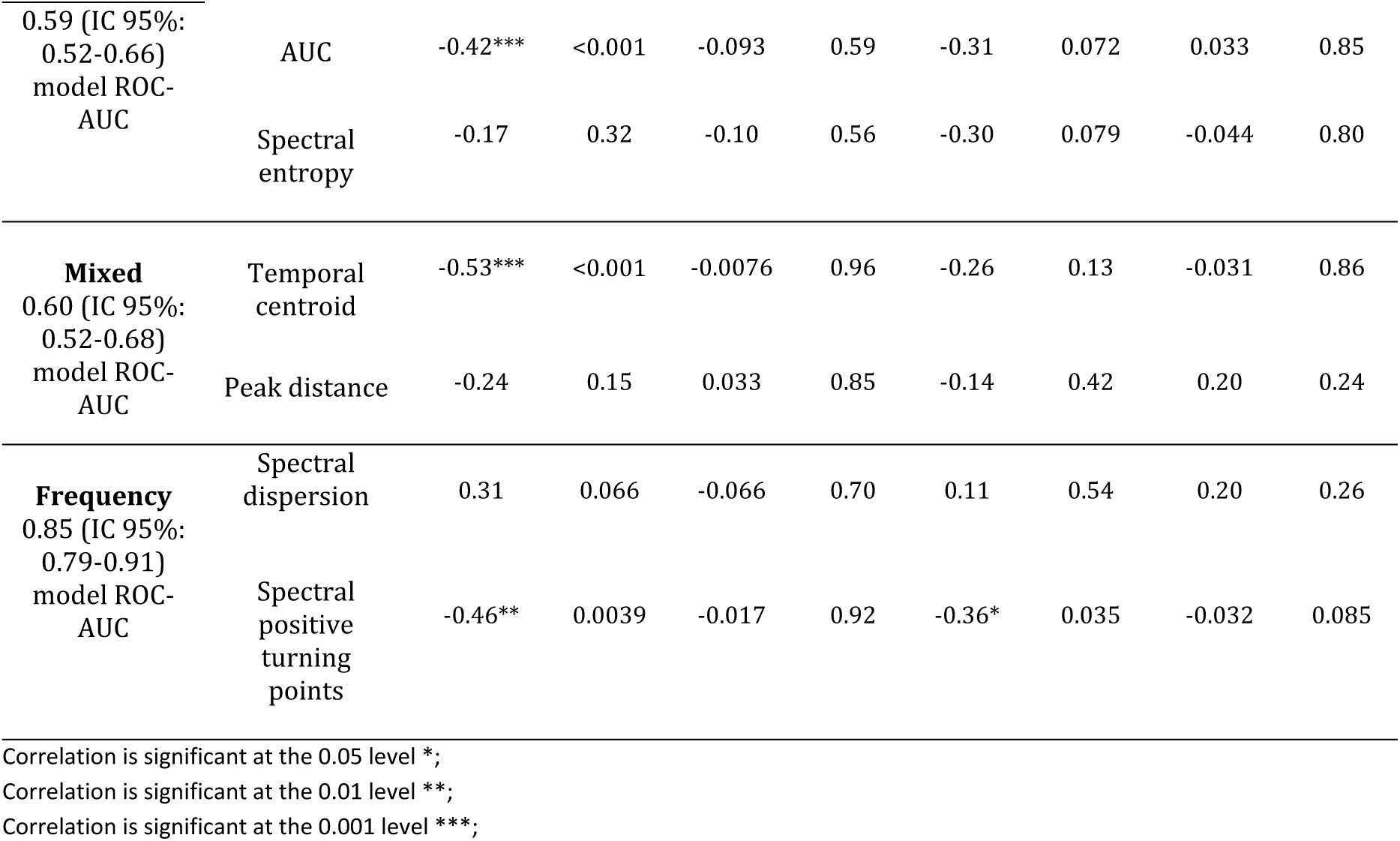
Correlations between the FVC (%), MIP (%), MEP (%), and CPF (L/min), and different cough sound features.

### 3.5 Differences between ALS patients with and without bulbar dysfunction

When comparing ALS patients with and without bulbar dysfunction, we observed that frequency-related features were the best group at this discrimination (**Table 7**). The frequency features that were retained in the final model included spectral bandwidth, spectral centroid, spectral roll-off, and spectral kurtosis and skewness. Despite being the most significant, the overall ROC-AUC of the model prediction was 0.53 (IC 95%: 0.44-0.61).

**Table 7.** F values from regression analyses contributing of with bulbar vs. without bulbar dysfunction classification to performance on each voice sound variable.

| GROUP | FEATURES | F value | P value |
| --- | --- | --- | --- |
| **Frequency group** | Spectral bandwidth | 6.07* | 0.017 |
|  | Spectral centroid | 11.76** | 0.0011 |
|  | Spectral roll-off | 4.79* | 0.033 |
|  | Spectral Kurtosis | 6.23* | 0.015 |
|  | Spectral Skewness | 8.44** | 0.0052 |
| **Intensity group** | Temporal centroid | 1.41 | 0.24 |
|  | Maximum amplitude of the signal | 2.15 | 0.15 |
|  | Mean amplitude of the signal | 1.35 | 0.25 |
|  | Median amplitude of the signal | 0.38 | 0.54 |
|  | Standard deviation of the signal amplitude | 0.16 | 0.70 |
|  | Variance amplitude of the signal | 0.27 | 0.61 |
|  | Peak-to-peak distance | 4.29* | 0.043 |
| **Mixed group** | Area under the curve of the signal | 0.83 | 0.37 |
|  | Maximum power of the signal | 3.86 | 0.054 |
|  | Spectral entropy | 0.66 | 0.42 |

Correlation is significant at the 0.05 level \*; Correlation is significant at the 0.01 level \*\*;

As for the intensity-related features, the predictors found in the final model included temporal centroid, maximum, mean, median, standard deviation, variance, and kurtosis of the signal. The final ROC-AUC of the model prediction for the intensity group was 0.63 (IC 95%: 0.51- 0.75).

Lastly, for the features related to the mixed group, the ones that remained in the model were the area under the curve of the signal, maximum power, spectral entropy, and gender. The final ROC-AUC of the model was 0.51 (IC 95%: 0.39-0.63).

## 4.0 Discussion

Our study aimed to comprehensive investigate the potential of cough sound features, extracted from both the time and frequency domains, as discriminators for clinical diagnosis of ALS, and predictors of bulbar and respiratory impairments, at the convenience of using a simple smartphone. Based on our hypothesis, significant differences were observed in the frequency group of features between ALS patients and healthy controls, after adjustment for age and gender. This was also the group of features that demonstrated higher correlations with bulbar impairments. Conversely, the intensity and mixed groups of features were found to be highly correlated with the functional status of the disease and were the most significant in detecting respiratory impairments (**Table 8**).

**Table 8.** Summary of all correlations undertaken in this study. The symbol ‘*check*’ denotes statistical significance, *α*=0.05 was considered.

|  |  | Control | Alsfrs-R | Bulbar | Respiratory | FVC% | MIP% | MEP% | CPF |
| --- | --- | --- | --- | --- | --- | --- | --- | --- | --- |
| **Frequency group** | Maximum frequency | ✓ | - | ✓ | - | - | - | - | - |
|  | ZCR | ✓ | - | - | ✓ | - | - | - | - |
|  | Spectral centroid | - | ✓ | ✓ | - | - | - | - | - |
|  | Sp. bandwidth | ✓ | ✓ | ✓ | - | - | - | - | - |
|  | Sp. dispersion | ✓ | - | - | - | - | - | - | - |
|  | Sp. Positive turning points | ✓ | - | - | - | ✓ | - | ✓ | - |
| **Intensity group** | Maximum amplitude | - | ✓ | - | ✓ | - | - | - | - |
|  | Peak distance | - | ✓ | - | ✓ | - | - | - | - |
|  | Temporal centroid | ✓ | - | - | - | ✓ | - | - | - |
| **Mixed group** | AUC | - | ✓ | - | - | ✓ | - | - | - |
|  | Maximum power | - | ✓ | - | ✓ | - | - | - | - |
|  | Sp. entropy | ✓ | - | ✓ | ✓ | - | - | - | - |

Firstly, changes in sound frequencies during any type of vocalization are primarily attributed to intrinsic modifications of the vocal cords. These variations in sound tone are intricately linked to the vocal cords’ dimension, tension, and/or thickness (37). In the present work, we noticed that the disease-related ALS cough is hoarser when compared to the controls – i.e., patients’ cough depicts lower frequencies (more specifically, lower zero-crossing rates and spectral positive turning points). These results suggest that the bulbar region of the glottis in ALS patients potentially exhibits increased tension and reduced flexibility, as higher levels of tension tend to produce lower frequency sounds (38–40).

Another finding, closely related to the previous, was that the cough produced by ALS patients displays greater sound entrainment, greater noise, and reduced sound occlusion when compared to controls. Occlusive sounds result from the obstruction or blockage of airflow in the vocal tract, and they are representative of functional cough sounds. In ALS, the adductor muscles of the arytenoid cartilages become dysfunctional (41), the glottis is not rapidly coordinated and fails to close effectively, leading to an abnormal compressive cough phase. Consequently, the typical peak in cough sound amplitude is not succeeded by a period of silence, but it is rather followed by an entrainment of the expiratory airflow. This was broadly represented by higher spectral dispersion and bandwidths. In fact, the cough sound properties of ALS patients resemble the characteristics of a sustainable vowel sound – a monophonic sound characterized by a continuous flow of air through the vocal cords. Moreover, as an alternative, the representation of the cough sound (presented in **Fig 3**) can also be explained by the varying properties of the medium through which the sound wave travels, such as different pressures and tensions, resulting in differing wave speeds and wave spread.

As a whole, the above-mentioned observations are in line with the evidence reported in cough airflow studies and cough waveforms visual analysis in patients with motor neuron diseases (42,43). Chaudri et al. (43) characterized the absence of distinct “peak expiratory spikes” and associated this with reduced cough strength and increased mortality. Recently, Plowman et al. (44) demonstrated that ALS patients showed lower peak expiratory flow rates and a longer time to generate maximum expiratory flow during a voluntary cough. They observed that this less efficient expulsive cough (as indexed by a lower cough volume acceleration) is predictive of poor airway safety during swallowing. Moreover, Korpáš et al. (45) have reported that, in laryngeal inflammation, the cough record consists of a large and long mono sound, where both sound intensity and duration may be increased. Thus, the cough sound in ALS may be associated with a secondary inflammation as well. All the aforementioned attributes collectively rendered this set of features superior in discriminating between cough sounds of patients and controls, resulting in a final model ROC-AUC of 0.85 (IC 95%: 0.79-0.91).

Emphasizing these primary distinctions in cough sounds between patients and controls, it is noteworthy that the observed differences between the two groups also manifested when comparing patients with and without bulbar symptoms. In this particular analysis, even though the machine-learning classifier did not exhibit exceptional robustness (with comparable results among frequency, intensity, and mixed feature groups), the most impactful features in distinguishing the two groups were maximum frequency, spectral bandwidth, and spectral centroid. Notably, these features are easily perceptible to the human ear, as healthy cough sounds typically display a clear quality, even when accounting for variations in age and gender. This enables clinicians to develop early suspicions regarding disease progression.

Furthermore, the intensity and mixed groups of features did not exhibit many significant differences between patients and healthy subjects. It is established that intense or louder sounds are related to higher air volumes in the lungs and consequently higher subglottic pressures. Despite 25 out of the 60 patients presenting respiratory dysfunction, as defined by scores less than 12 in the three respiratory-related questions of the ALSFRS- R (although many patients only presented with one less point) and a moderate to low FVC in the population, we speculate that these characteristics were insufficient to detect changes in intensity features, such as signal amplitude or peak distance, when compared to the cough sound of controls. Additionally, bulbar impairments such as a narrowed glottis are more likely to become clinically symptomatic when respiratory muscles are still strong enough to generate negative airway pressure(46). Nonetheless, patients exhibited higher spectral sound entropies and temporal centroids, meaning that the cough sounds are more variable and difficult to predict, and the average energy of the sound, occurs later in time (also related to wave spread and power). For these reasons, the sets of intensity and mixed features exhibited the lowest level of ROC-AUC, with the latter outperforming the former (ROC-AUC of 0.62, 95% CI: 0.55-0.69 and 0.70, 95% CI: 0.64-0.76, respectively), primarily due to the presence of features associated with sound frequency.

To verify the relationships between cough sound features and the respiratory system, the same approach was utilized, first to assess correlations with variables from respiratory function tests, and second to evaluate differences in patients with and without respiratory dysfunctions.

In ALS patients, coughing is impaired during both the inspiratory and expiratory phases, with lower volumes of inspired air during a prolonged inspiratory phase and a longer time period to generate a lower peak expiratory airflow during the expulsive phase (as reported by (41)). Additionally, studies have demonstrated that the volume of air achieved at the initiation of the cough has the greatest influence on the volume expelled during cough (47). In sound analysis, loudness and intense sounds are related to volume. This relationship further reinforces the significance of the intensity-based cough sound features as the most reliable indicators of respiratory impairment in patients. Furthermore, the cough sound pattern exhibited by ALS patients is consistent with that observed in patients with restrictive respiratory disease, characterized by reduced lung elasticity or limitations in chest wall expansion (16). In these patients, there is a gradual reduction in the intensity of cough attempts over time, leading to a negative slope of the signal amplitude (**Fig 4**). This is in contrast to obstructive respiratory diseases, where such a phenomenon is not observed. In the machine learning analysis, the model that demonstrated the highest ROC-AUC (although also not particularly robust overall), in distinguishing between patients with and without respiratory symptoms was the one trained with intensity-related features (0.67; IC 95%: 0.56-0.78). However, and despite these findings, the exact role of different respiratory muscles and their association with these cough sound features remains unclear. To understand this relation, we performed linear regressions between the cough sound features and FVC% values. FVC is highly associated with CPF measurements in ALS patients (Matsuda et al. 2019). Sharan et al. (12), demonstrated the potential for cough sound analysis to predict spirometry results in patients with different respiratory diseases. In this work, the intensity and mixed group of features, specifically the temporal centroid and absolute energy, exhibited stronger correlations with FVC%. These findings provide support for the association between sound energy, intensity, and lung function. Notably, the correlations between FVC% and energy features are negative. This finding may indicate that patients with respiratory dysfunctions often experienced increased efforts to move air in and out of the lungs and even that, as it becomes difficult to fully exhale air, leading to air trapping, the trapped air during the subsequent cough bouts has contributed to higher sound energies. It was also anticipated that stronger correlations would be observed between cough sound features and MEP%, in comparison to MIP%, given that pulmonary exhalation is the primary source of energy for sound production.

MEP represents the highest achievable pressure during forceful expiration against a closed airway and indicates the strength of the abdominal muscles and other expiratory muscles. Conversely, MIP assesses the strength of inspiratory muscles, primarily the diaphragm, and enables the evaluation of ventilatory insufficiency. Although only the spectral positive turning points showed a significant correlation with MEP%, sound energy exhibited the potential to serve as a valuable distinguishing feature as well. Moreover, no significant associations were observed between cough sound features and CPF. This test involves coughing forcefully into a face mask connected to a small peak flow meter, and it measures the expelled airflow. We speculate that its precision may be limited by acoustic variations, particularly considering that cough sounds were captured laterally from the mouth, rather than directly by the smartphone microphone, to mitigate interference from wind noise. This represents a distinct analytic approach.

Some limitations of this study must be acknowledged. Specifically, voluntary coughs bypass the sensory system and previous research has demonstrated that maximum voluntary cough function tends to overestimate reflexive cough function among healthy volunteers (47,48). Moreover, the current study includes patients with mild-moderate disease severity. As a result, the generalization of these findings to airway defense in the event of aspiration as well as to individuals in a more advanced disease state may be limited. Further, given the clinical heterogeneity of ALS, it would be beneficial to document upper versus lower motor neuron involvement, and slow versus fast progress to develop more homogenous groups for comparison. It is also possible that more appropriate features (as well as other machine learning models) may be extracted from the data, even when features that do not contribute to the model prediction ROC-AUC were eliminated. Performing a longitudinal cough sound analysis, recording cough sounds in a lying position, making clinical correlations with phrenic nerve conduction measures and muscle strength of cervical muscles, and adjusting the results for other motor neuron diseases are future perspectives that could help elucidate the results of this paper.

## 5.0 Conclusion

The present study demonstrates that analyzing cough sounds can serve as a valuable technique for evaluating and monitoring ALS patients, particularly those with respiratory and bulbar impairments. However, it is important to note that cough sound analysis should not be the only indicator utilized to evaluate respiratory and bulbar health, as ALS is a multifaceted and intricate disease. Rather, it can be used as an adjunct measure, supplementing commonly used ways of disease progression. It is also noteworthy that the method used in this study was a convenient smartphone-based approach, which facilitates data collection in home-based settings without requiring specialized careers or equipment.

## Data Availability

All relevant data are within the manuscript and its Supporting Information files.

## Financial Disclosure Statement

This study was part of a broader ALS project (HomeSenseALS - PTDC/MEC-NEU/6855/2020), supported by the Foundation for Science and Technology.

## Competing interest

No potential conflict of interest was reported by the author(s).

## Supporting information

**S1 File. Dataset encompassing all the information used for comparing a cough sound associated with ALS and a cough sound from a healthy control.**

**S2 File. Dataset encompassing all the information used for comparing cough sound features and clinical variables from ALS patients.**

## Notes

### Competing Interest Statement

The authors have declared no competing interest.

### Funding Statement

Yes

### Author Declarations

The study was approved by the local research ethics committee of the Centro Académico de Medicina de Lisboa (CAML-Ref. 146/21).

